# Natural immune boosting can cause synchrony in susceptibility and outbreaks of respiratory infections with rapidly waning immunity

**DOI:** 10.1101/2023.11.23.23298952

**Authors:** Mark G. Pritchard, Sean M. Cavany, Susanna J. Dunachie, Graham F. Medley, Lance Turtle, Christl A. Donnelly, Peter W. Horby, Ben S. Cooper

## Abstract

Natural immune boosting is a mechanism in which re-exposure to a pathogen while immunity is waning can prolong protection against reinfection. Its role in viral respiratory-tract infections with rapidly waning immunity has not been studied in mathematical models. Here we use a deterministic compartmental model to investigate the effect of immune boosting on such infections, and how the reduction in transmission due to non-pharmaceutical interventions during the covid-19 pandemic could affect immune waning and boosting. We find that immune boosting can introduce unstable equilibria into the model, and we show in simulations that this can amplify, or even cause, oscillations in infections and disease outbreaks. After periods of reduced transmissibility, representing non-pharmaceutical interventions, simulations with more immune boosting exhibit larger rebound outbreaks that occurred sooner. Observed incidence of respiratory syncytial virus infection in Scotland between 2016 and 2023 can be equally explained by models with high levels of immune boosting, and models without natural immune boosting. To produce the same incidence, models with more immune boosting require a greater mean transmissibility, suggesting that models underestimating natural immune boosting will also underestimate transmissibility.

---

Natural immune boosting is a mechanism first proposed by Hope-Simpson [1] in which re-exposure to a pathogen while immunity is waning prolongs protection against disease. Such boosting is important for prolonging immunity to pertussis [2], measles and mumps [3], and for reducing risk of severe disease from malaria [4] and covid-19 [5]. Epidemiological compartmental models with susceptible–infectious–recovered–susceptible (SIRS) dynamics can be modified to account for natural immune boosting by adding one or more categories of individuals with partially waned immunity [6–11]. These individuals either continue waning to susceptibility or return to the fully-immune category, the latter occurring at a rate proportional to the force of infection (the probability of a waning individual’s immunity being boosted is proportional to the probability of a susceptible individual becoming infected).

Viral infections of mucosa, including the respiratory tract, elicit briefer immunity than systemic infections [12]. Reinfection with respiratory viruses can occur across all ages and, in contrast to endemic ‘childhood’ diseases, severe disease is common among the elderly [13]. Even while systemic protection against severe disease persists, mucosal immunity can wane sufficiently for infectious virus to be present in respiratory secretions [14]. Many respiratory viruses display seasonal behaviour at both temperate and tropical latitudes, which may be only partially explained by seasonal environmental changes and patterns of social interaction [15–18]. Reinforcing vaccination is recommended to boost immunity for individuals at high risk of severe disease from influenza virus and severe acute respiratory syndrome coronavirus 2 (SARS-CoV-2). The effect of immune boosting by natural re-exposure to respiratory viruses has not been widely studied and has not been the subject of mathematical models.

Respiratory syncytial virus (RSV) infection elicits remarkably brief immunity. Rein-fection is possible within two months [19], and each year 4–10% of adults are reinfected [20]. In 2020, many governments mandated non-pharmaceutical interventions to reduce the spread of SARS-CoV-2, such as school closures, restrictions on gathering, and requirements to wear face coverings [21]. This disrupted RSV transmission, and the typical annual outbreaks were not seen in 2020 [22–24]. Unusually early and large outbreaks of RSV occurred in the following two years, attributed to increased susceptibility in the population [25–27].

Here we use a deterministic compartmental model to investigate the effect of immune boosting on respiratory virus infections with rapidly waning immunity. We focus on disease dynamics so define immunity in our model as protection against becoming infectious. We assume that boosting returns individuals to full immunity without causing infectiousness. A boosting coefficient, *ψ*, is defined as the ratio of boosting to the force of infection, such that *ψ* = 0 represents no immune boosting and *ψ >* 1 implies that boosting occurs more readily among those with waning immunity than infection does among those susceptible. Large values of the boosting coefficient, such as *ψ ≥* 10, have been considered plausible in the context of pertussis [6, 7]. Compartmental models with immune boosting have not previously considered pathogens with rapidly waning immunity. We analyse the model’s equilibrium points for a range of potential boosting coefficient values and explore the underlying effects of immune boosting on disease dynamics. We then use parameters that mimic reported RSV infections in Scotland between 2016 and 2023 to consider how immune boosting could affect transmission, both in pre-covid ‘steady state’ conditions and after the introduction of non-pharmaceutical interventions.

## Results

We assumed SIRS-like dynamics with two additional compartments for individuals with partially waned immunity (Figure 1; see *Methods* for full model details). The model’s disease-free equilibrium, *S^∗^* = 1, is stable if and only if the basic reproduction number, *R*_0_ *<* 1. The endemic equilibrium proportion susceptible is 1*/R*_0_ when *R*_0_ *>* 1 so is independent of waning and boosting parameters (Figure S1). Without immune boosting, the model follows SIRS dynamics, with the serial recovered subcompartments giving an Erlang-distributed duration of immunity (Figure S2). In this case, the equilibrium proportion infectious increases monotonically with greater values of *R*_0_ (when *ψ* = 0 in Figure S1*B*). With immune boosting, *ψ >* 0, there is a turning point beyond which greater infectiousness is associated with smaller equilibrium proportions infectious and larger proportions immune.

**Figure 1:**
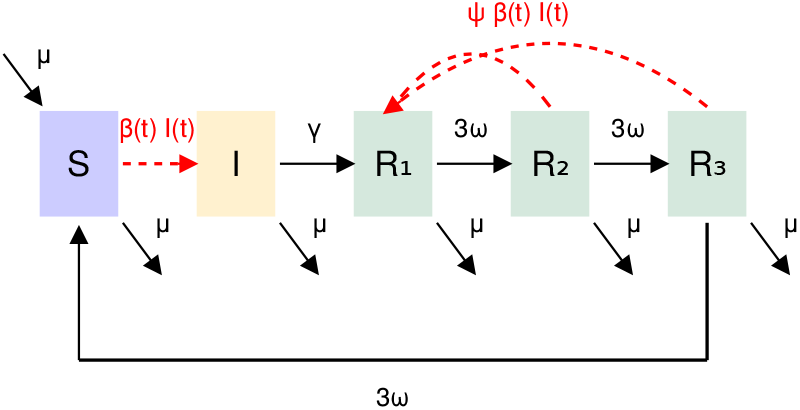
Model flow chart. Compartments are, *S*, susceptible; *I*, infectious; *R*_1_–*R*_3_, resistant (immune). Dashed red lines represent events that occur in proportion to the force of infection. See *Methods* for parameter definitions.

With no immune boosting, these stationary points are stable. Greater values of the boosting coefficient lead to a Hopf bifurcation beyond which equilibria are unstable. We denote the threshold boosting coefficient value that separates stable and unstable equilibria as *ψ^∗^*. For a pathogen eliciting immunity that would last approximately two years without boosting, we found low values of *ψ^∗^*, suggesting that boosting at a similar level of exposure as required for infection could lead to instability (Figure 2*A*). The magnitude of *ψ^∗^* increases rapidly as the unboosted immune duration approaches zero, meaning that as the model approaches susceptible–infectious–susceptible dynamics, equilibria are stable even if immune boosting requires much less exposure than infection. When the boosting coefficient is greater than *ψ^∗^*, compartment sizes do not converge on the calculated equilibrium values. Instead they approach a limit cycle, oscillating between a larger and a smaller proportion infectious (Figure 2*B* and *C*, and Figure S3). The interval between outbreaks increases (reduced frequency) with greater boosting coefficients (Figure S4*A*).

**Figure 2:**
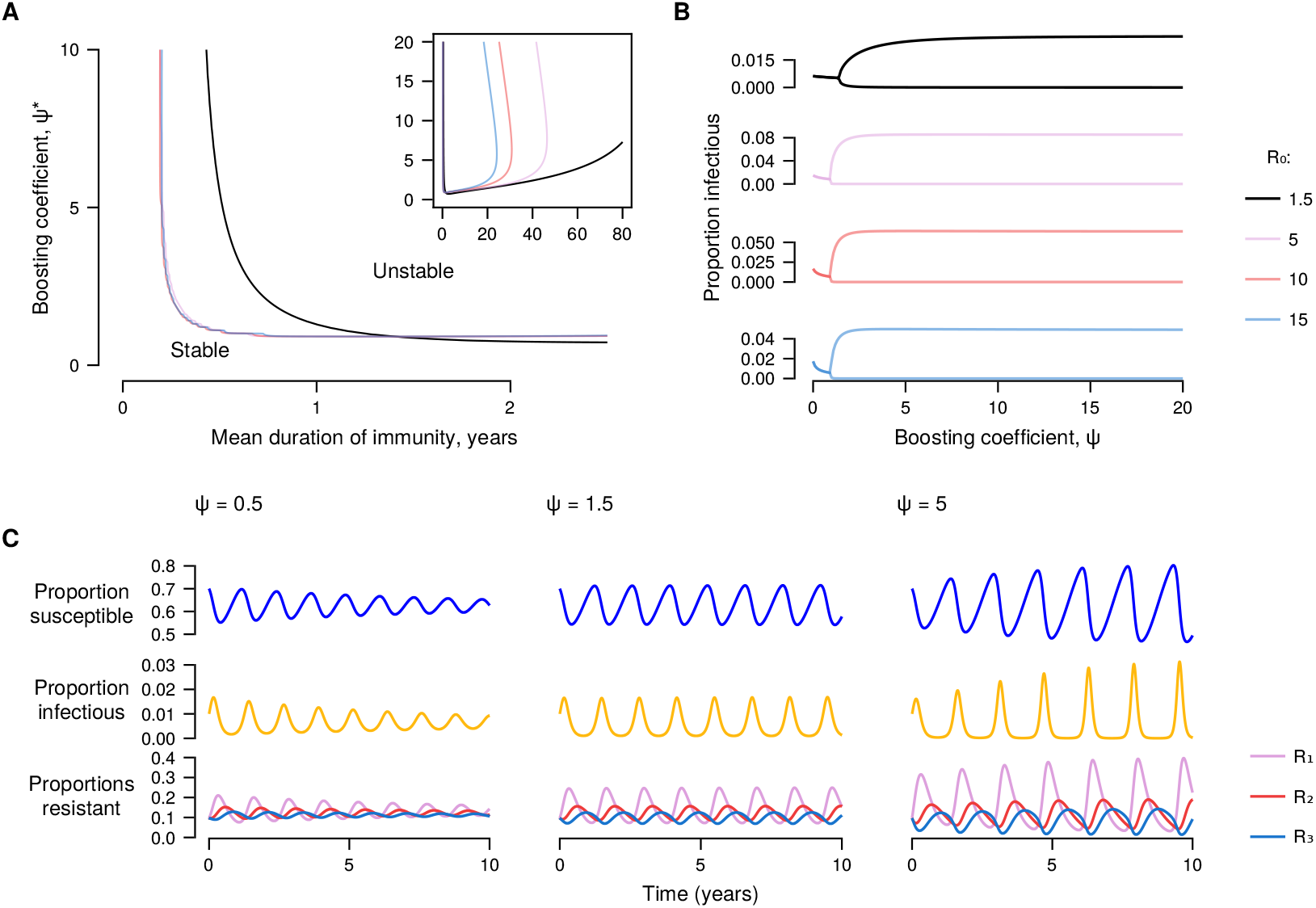
Effect of boosting coefficient on equilibrium stability. (*A*) Threshold values of the boosting coefficient, *ψ^∗^*, for the Hopf bifurcation separating stable from unstable equilibria. Inset shows thresholds for longer durations of immunity. (*B* ) Bifurcation plots showing maximum and minimum proportions infectious. Line colours in *A* and *B* indicate *R*_0_ values. (*C* ) Simulated proportions in each compartment, with *R*_0_ = 1.6 and *ψ^∗^* = 1.18. Recovered subcompartments *R*_1_ to *R*_3_ are entered in order, with *R*_1_ representing those with the greatest expected time before returning to susceptibility. In each plot, birth rate is 0.087 per year, mean generation time is 7.5 days, and mean immune duration without boosting is 400 days.

We simulated seasonal forcing of transmission by varying the transmission parameter sinusoidally over the year. We were able to generate qualitatively indistinguishable patterns of incidence in simulations with seasonal forcing and no immune boosting, and in simulations with no seasonal forcing and a large boosting coefficient (see first five years of simulations in Figure 3). With intermediate values, if the frequency of seasonal forcing was close to the resonant frequency of the unforced system then stable annual outbreaks were seen (see strong signals correlated with annual or two-yearly outbreaks for a range of boosting coefficients in Figure S4*B*). Simulations with more immune boosting required greater mean transmissibility to generate equivalent incidence patterns. Greater immune boosting led to greater proportions in the first resistant subcompartment during each outbreak. This was followed by a rapid, synchronized return to susceptibility between outbreaks. As expected for models with larger *R*_0_, the proportion susceptible was smaller in simulations with more immune boosting.

**Figure 3:**
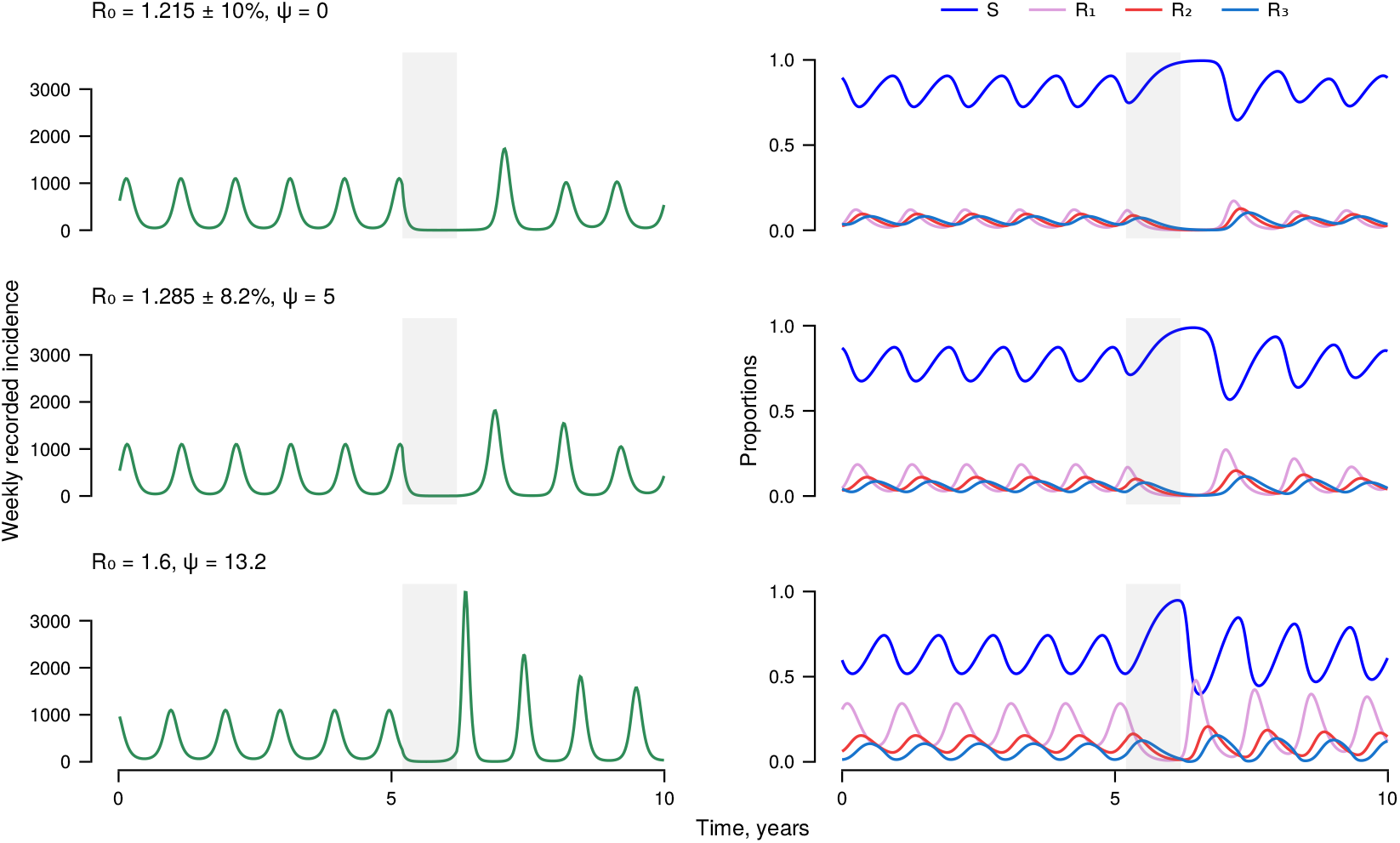
Simulated incidence, before and following non-pharmaceutical interventions. Left-hand plots show simulated weekly incidence assuming a population of 5.5 million and detection of 1% of infections. Right-hand plots show compartment sizes. Grey shaded area represents a period with 20% reduction in transmission. *±* values represent amplitude of transmission seasonal forcing. In each plot, birth rate is 0.087 per year, and mean generation time is 7.5 days.

The similarity between these simulations suggested that unique model parameters might not be identifiable, even if incidence data could be recorded without error. We investigated the model’s structural identifiability using Castro and de Boer’s [28] method. Each model parameter was multiplied by an unknown scaling parameter. If any of these scaling parameters could take values other than 1 then the model parameters were not uniquely identifiable. We found that, given prevalence data (i.e. observing *I*(*t*)), the immune waning parameter and proportions in recovered subcompartments could not be uniquely identified (see *Supplementary text* ) so the model parameters were nonidentifiable. We therefore explored the effect of a range of parameter values on the dynamics of immunity and disease transmission.

We simulated adoption of non-pharmaceutical interventions by transiently reducing the transmission parameter (shaded grey area in Figure 3). In all simulations, nonpharmaceutical interventions led to a reduction in incidence and an increase in the proportion susceptible. Once non-pharmaceutical interventions were lifted, simulations with the greatest boosting coefficient (and the greatest *R*_0_) exhibited larger rebound outbreaks that occurred sooner.

Between 2016 and 2019, RSV diagnoses in Scotland peaked each year between November and January (Figure 4). There was a secular trend of increasing numbers of diagnoses each year, from 3100 to almost 5000 (to avoid separating outbreaks across calendar years, we counted each year’s cases from April 1 onwards). In the twelve months from April 2020, 61 RSV infections were reported. In 2021, case numbers started to increase in August, leading to an earlier peak in RSV diagnoses but a total number similar to prepandemic years (3581 cases). In 2022, reports started in June and the total number of cases was 7331. Results stratified by age group (Figure S5) showed greater numbers of diagnoses in 2022 for all age groups, suggesting that reinfection was an important contributor to the size of that year’s outbreak.

**Figure 4:**
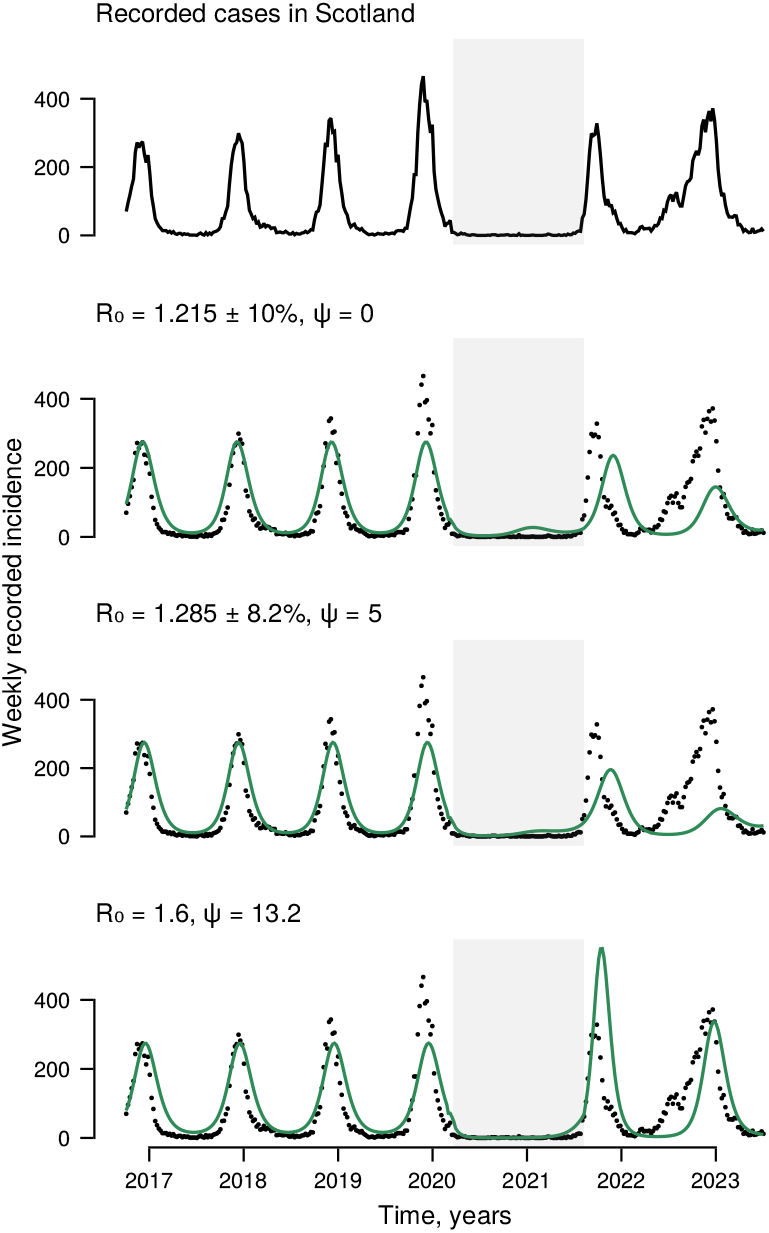
Respiratory syncytial virus in Scotland. The top plot, and black points in subsequent plots, show recorded respiratory syncytial virus infections in Scotland. Green lines represent simulations with different boosting coefficient values. Grey shaded area represents the period of high stringency index (*>*50) in Scotland. In each plot, birth rate is 0.087 per year, and mean generation time is 7.5 days.

Assuming that, before non-pharmaceutical interventions, all infants were infected in the first two years of life [29] and 4–10% of adults were reinfected annually [20], at least 98% of RSV infections were unrecorded. We selected parameters that gave qualitatively good fits to the data, assuming that 0.25% of infections were recorded (Figure 4). We assumed a reduction in transmission in Scotland between March 21, 2020 and August 8, 2021, corresponding to a period of high stringency on the Oxford Covid-19 Government Response Tracker [21] (Figure S6). We found that assumptions about immune boosting altered model-derived estimates of the effect of non-pharmaceutical interventions. With no immune boosting, a 10% reduction in transmission minimized the error between the simulation and RSV incidence. If we assumed a high level of immune boosting, *ψ* = 13.2, then the estimated effect of non-pharmaceutical interventions was much greater, 26%.

## Discussion

Our results suggest that natural immune boosting to viral respiratory-tract infections with rapidly waning immunity could lead to synchrony in population immunity and susceptibility. With sufficient boosting, this could cause periodic peaks of infection even with no seasonal changes in transmissibility. At lower levels of immune boosting, these inherent oscillations can resonate with, and amplify, the effects of seasonal changes. We demonstrated that observed patterns of RSV infections before and after the covid-19 pandemic, could be consistent with a wide range of boosting coefficients. This included situations in which no natural immune boosting occurs, and in which boosting among those whose immunity is waning occurs at 13 times the rate of infection among those who are susceptible. Mischaracterization of immune boosting can lead to errors in model-derived estimates of other parameters. If the effect of immune boosting is assumed to be lower than it is, then we will underestimate the pathogen’s basic reproduction number, and underestimate the magnitude of reduction in transmission required to control spread.

Synchronous and oscillatory dynamics are widely recognised in epidemiology [30], and more broadly across biological processes [31]. Even when disease dynamics would tend toward stability, if periodic environmental effects resonate with the system’s intrinsic oscillations, this can lead to large seasonal variation in the incidence of infection [32]. In systems without immune boosting, more frequent outbreaks are predicted when susceptible numbers are rapidly replenished by high birth rates [33, 34] or short durations of immunity [32]. If these rates are non-constant then outbreaks may synchronize with the susceptible replenishment. For instance, seasonality of animal birth rates contributes to oscillatory dynamics of their diseases [35]. In our model, the effective rate of immune waning is inversely related to the prevalence of infection (boosting prolongs immunity). Immune boosting reduces the return to susceptibility during outbreaks. These outbreaks are followed by more rapid immune waning and synchrony in the return to susceptibility, priming the population for another large outbreak. Dafilis and colleagues [8] found that increasing life expectancy in a compartmental model of boosting immunity to pertussis led to oscillatory dynamics. In these models, increasing life expectancy (reducing mortality rate) equivalently reduces the birth rate. This increases the relative contribution of immune waning to the size of the susceptible compartment, in a similar way to increasing the waning rate (reducing the duration of immunity) in our model.

Oscillatory behaviour in disease dynamics may be desirable if it is sufficiently extreme to cause local elimination of a pathogen [30]. However, for viruses such as RSV with rapidly waning immunity and globally high prevalence, reintroduction is expected to occur so readily that local elimination is implausible [36]. Rather than benefiting from oscillations, large peaks in numbers infected may overwhelm healthcare systems, and public health authorities may instead need to direct efforts to ‘flattening the curve’ [37]. We have not included vaccination in our model, but the potential for an outbreak driven by the return to susceptibility suggests a possible strategy for repeat vaccination to be timed with the nadir of the force of infection. Explicit consideration of the balance between the effects of immune waning and seasonal forcing would be needed to determine the optimal vaccination strategy.

Considering RSV specifically, the monoclonal antibody nirsevimab and several vaccines have recently been licensed, or soon will be [38]. The global need to reduce the burden of RSV infection is clear. It caused an estimated 100 000 childhood deaths in 2019 [39] and causes up to a quarter of excess winter deaths among adults [13]. Modelling [40, 41] suggests that both would be cost effective. These models did not include the potential for natural immune boosting. If a widespread intervention among infants reduced exposure to the virus among adults, this could potentially cause a paradoxical increase in infection due to reduced immune boosting, similar to that reported for measles and mumps [3]. RSV tends to be under-diagnosed among adults, including those with severe disease, due to its non-specific symptoms [42]. Systematic surveillance for RSV infection among adults admitted to hospital with acute respiratory-tract infections may be needed to identify any unintended consequences of reducing transmission among children. Such surveillance should start before any vaccination programme.

Our analysis has many simplifying assumptions. We assumed all-or-nothing immunity with those whose immunity had waned being indistinguishable from those who were immune-näıve. This does not reflect realistic patterns of immunity for RSV, influenza [43], or SARS-CoV-2 [44]. Our model also neglects clinically-important protection against severe disease and death, which persists after mucosal immunity to infection has largely waned [45]. We assumed that the effects of starting and stopping non-pharmaceutical interventions were immediate, but during the covid-19 pandemic some restrictions remained in place for longer than others [21], and levels of engagement with restrictions varied over time [46]. Our model differs from many RSV-specific models that include distinct sequential susceptible, infectious and immune states to represent accumulating effects of re-exposure [47]. We assumed the pathogen was homogeneous, with no variation in strains and no evolution over time. Even for RSV, in which the immunogenic F protein is highly conserved [48], this may not accurately capture all relevant effects, as the dominant RSV group varies between outbreaks [49]. In models with SIRS dynamics, loss of immunity due to waning is equivalent to loss of immunity due to pathogen evolution [50]. Given that exposure to new strains of influenza virus can boost immune response to multiple previous strains [43], it may be plausible that a similar equivalence exists for our model of immune boosting. Alternatively, if a new variant caused infection rather than boosting immunity to a previous variant then different mechanisms of immune loss would lead to different patterns of immunity.

The wide range of plausible boosting coefficient values in our model replicates general uncertainty about many aspects of RSV transmission. White and colleagues [51] were able to fit dramatically contrasting models to RSV datasets, including one with lifelong partial immunity and *R*_0_ = 9.3, and another with expected immunity lasting less than 7 months and *R*_0_ = 1.3. Baker and colleagues’ [52, 53] model of the effects of non-pharmaceutical interventions on transmission produced qualitatively equivalent results to our model, but assumed lifelong immunity and *R*_0_ *≈* 7. Each model specification and set of parameters can lead to different predictions for disease dynamics. Underestimating natural immune boosting leads to underestimates of transmissibility and overestimates of the effect on incidence of a reduction in transmission. It is therefore vital to establish more robust understanding of natural immune boosting for respiratory-tract infections to inform successful disease control interventions.

## Methods

### Respiratory syncytial virus diagnosis data

Weekly counts of laboratory-confirmed RSV diagnoses in Scotland are publicly available [54]. We downloaded files of total and age-stratified diagnoses on July 18, 2023.

### The model

Our model assumed a constant population of 1 so that *S*(*t*), *I*(*t*) and *R*(*t*) were proportions susceptible, infectious and recovered (immune). A series of three recovered subcompartments, *R*(*t*) = *R*_1_(*t*) + *R*_2_(*t*) + *R*_3_(*t*), represent waning immunity. Our model is described by ordinary differential equations,

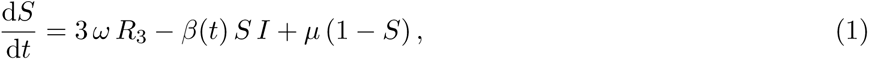

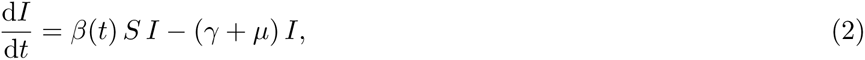

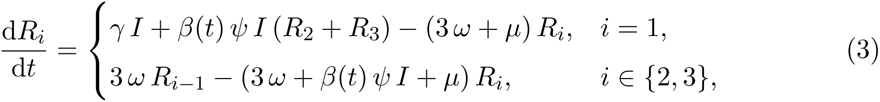

where 1*/γ* was the pathogen’s mean generation time, *µ* was the birth and death rate, and *ω* was the immune waning rate. To allow transmission to vary seasonally, we set,

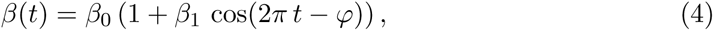

where *β*_0_ was the mean transmission parameter, *β*_1_ *∈* [0, 1] was the proportional amplitude of seasonal forcing, and *φ ∈* [0, 2*π*) was an offset for the time of maximum transmission. Simulation time was measured in years and all parameters were non-negative.

To find equilibria without seasonal forcing (*β*_1_ = 0, and *β*(*t*) is a constant *β*) we set equations 1–3 to equal 0. This was solved by disease-free conditions, 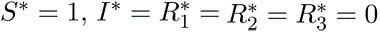, and by endemic equilibrium values,

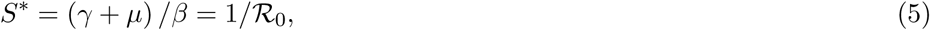

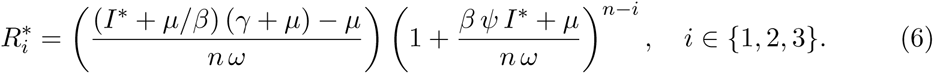

We solved equations 5 and 6 numerically for 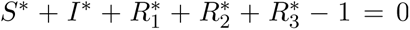, with all 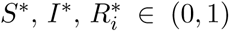. To assess stability we calculated eigenvalues of the model’s Jacobian at equilibrium. A Hopf bifurcation occurs if the real part of any eigenvalue becomes positive, then the equilibrium becomes unstable [31]. Eigenvalues at disease-free equilibrium were Λ_DFE_ = *{−µ, β − γ − µ, −*3 *ω − µ}*. Eigenvalues for endemic equilibria were calculated numerically in Julia, version 1.9.3 [55].

We used Castro and de Boer’s [28] scaling method to determine identifiability of parameters, assuming prevalence, *I*(*t*), could be observed without noise. See *Supplementary text* for details of the calculations.

### Simulations

Numeric simulations were performed using Verner’s [56] 9th order Runge– Kutta method in DifferentialEquations.jl, version 7.11.0 [57]. We set *µ* = 0.0087, approximating Scotland’s annual birth rate [58], and assumed a mean generation time of 7.5 days. Limit cycle values and discrete Fourier transforms were estimated from simulations with 1000-year warm-up periods, the latter using the fast Fourier transform algorithm in FFTW.jl, version 1.7.1 [59]. We estimated effects of non-pharmaceutical interventions by minimizing the squared error between the simulation and RSV incidence. We used Optimization.jl, version 3.19.3 [60], to estimate the effect of nonpharmaceutical interventions up until the stringency index was less than 50. We estimated the subsequent transmission through a series of iterations, using the previous estimate as the starting point for each following iteration.

### Ethical considerations

This analysis used publicly available data that did not allow potential identification of individuals. Ethics committee approval was not required.

### Software and code

Code and data to reproduce this analysis are available at https: //github.com/markgpritchard/ImmuneBoostingODEs. We used DrWatson.jl, version 2.13.0 [61] to manage a reproducible environment for all analyses. Figures were plotted using Makie.jl, version 0.19.11 [62].

## Data Availability

Code and data to reproduce this analysis are available at https://github.com/markgpritchard/ImmuneBoostingODEs

## Acknowledgments and funding sources

This study is funded by the National Institute for Health Research (NIHR) Health Protection Research Unit in Emerging and Zoonotic Infections (200907), a partnership between the United Kingdom Health Security Agency (UKHSA), the University of Liverpool, the University of Oxford and the Liverpool School of Tropical Medicine. The views expressed are those of the authors and not necessarily those of the NIHR, the UKHSA or the Department of Health and Social Care.

Data used in this analysis were released by Public Health Scotland under the Open Government Licence for public sector information, https://www.nationalarchives.g ov.uk/doc/open-government-licence/version/3/

## Author contributions

**M. G. Pritchard**, conceptualization, data curation, formal analysis, investigation, methodology, software, visualization, writing (original draft, review and editing); **S. M. Cavany**, formal analysis, methodology, writing (review and editing); **S. J. Dunachie**, writing (review and editing); **G. F. Medley**, writing (review and editing); **L. Turtle**, supervision, writing (review and editing); **C. A. Donnelly**, supervision, writing (review and editing); **P. W. Horby**, conceptualization, funding acquisition, supervision, writing (review and editing); **B. S. Cooper**, conceptualization, funding acquisition, methodology, supervision, writing (review and editing).

## Supplementary figures

**Figure S1:**
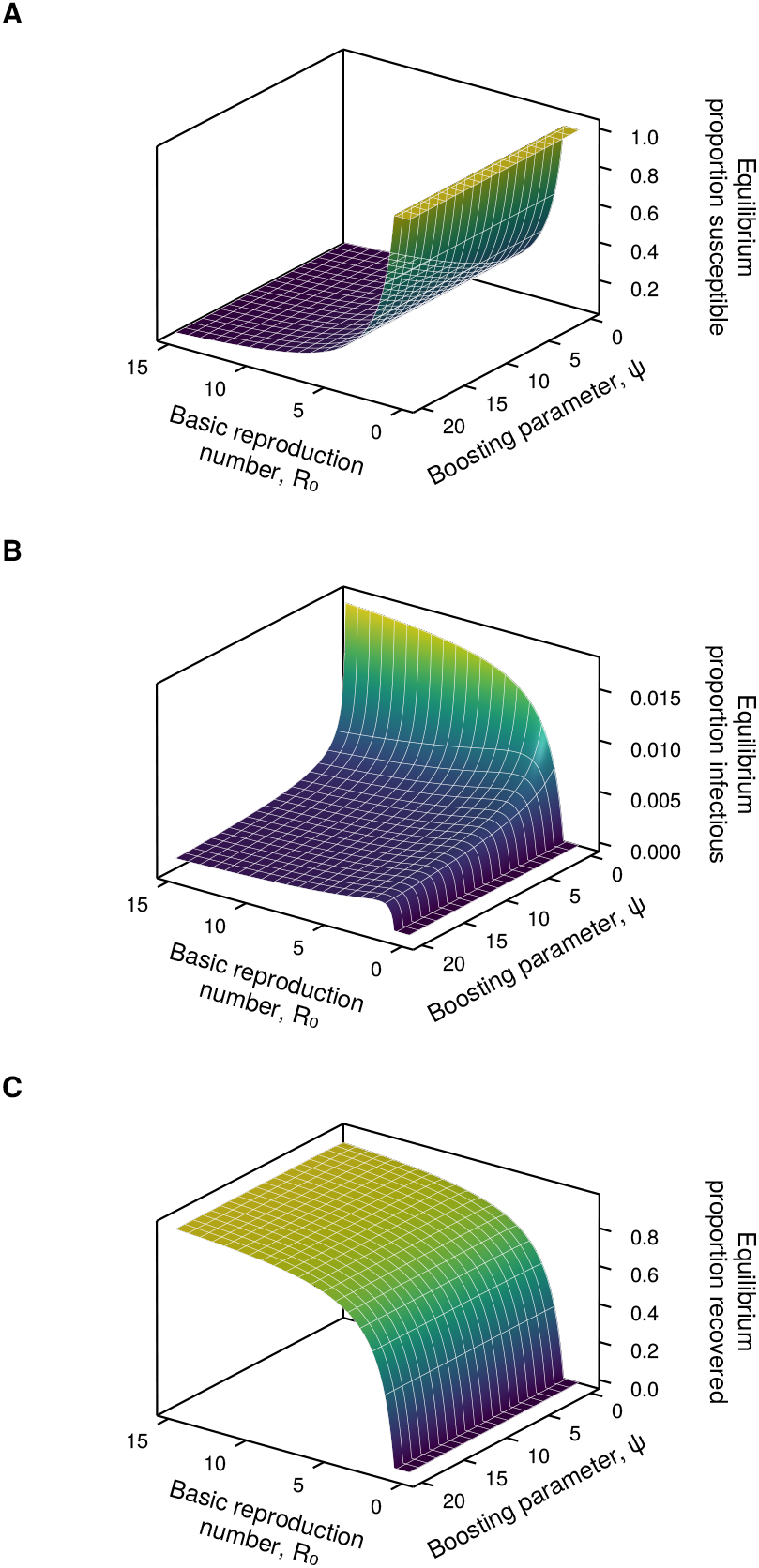
Proportions in each model compartment at calculated equilibrium. (A) Susceptible, (B) infectious, and (C) recovered (immune).

**Figure S2:**
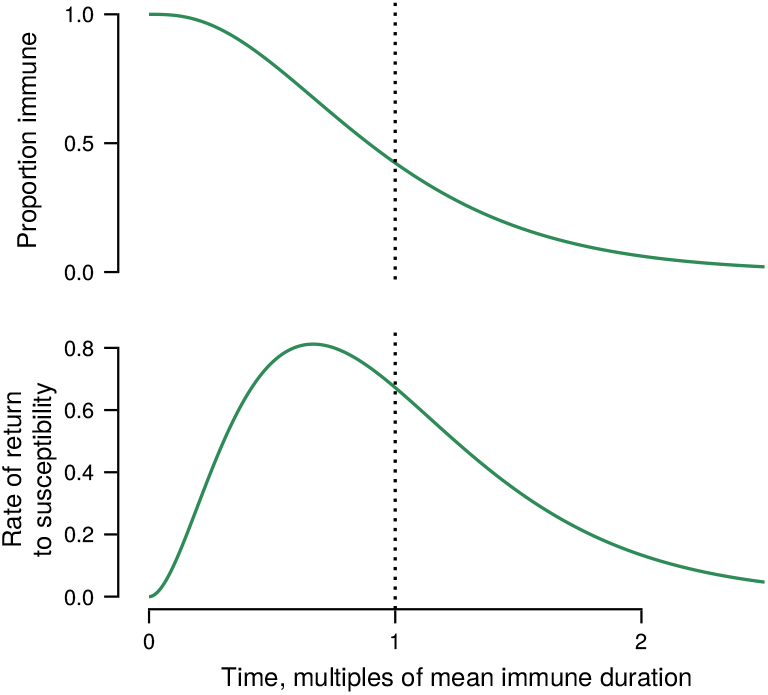
Model immune durations without immune boosting.

**Figure S3:**
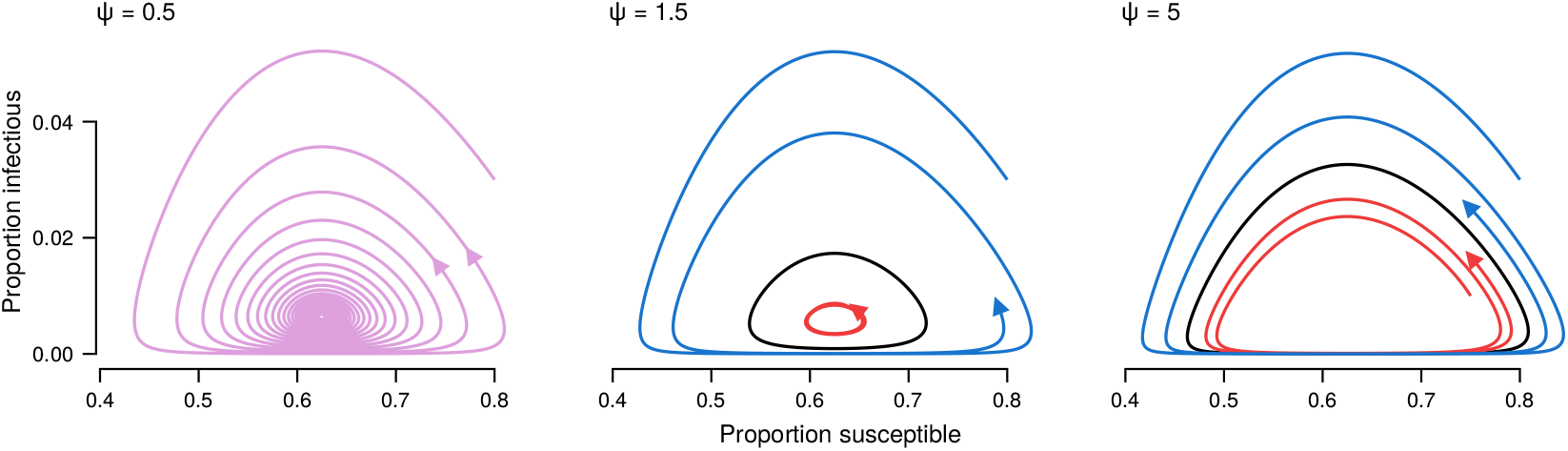
Infectious-susceptible planes for simulations with [inline] and *ψ*∗ = 1.18. Black loops indicate limit cycles.

**Figure S4:**
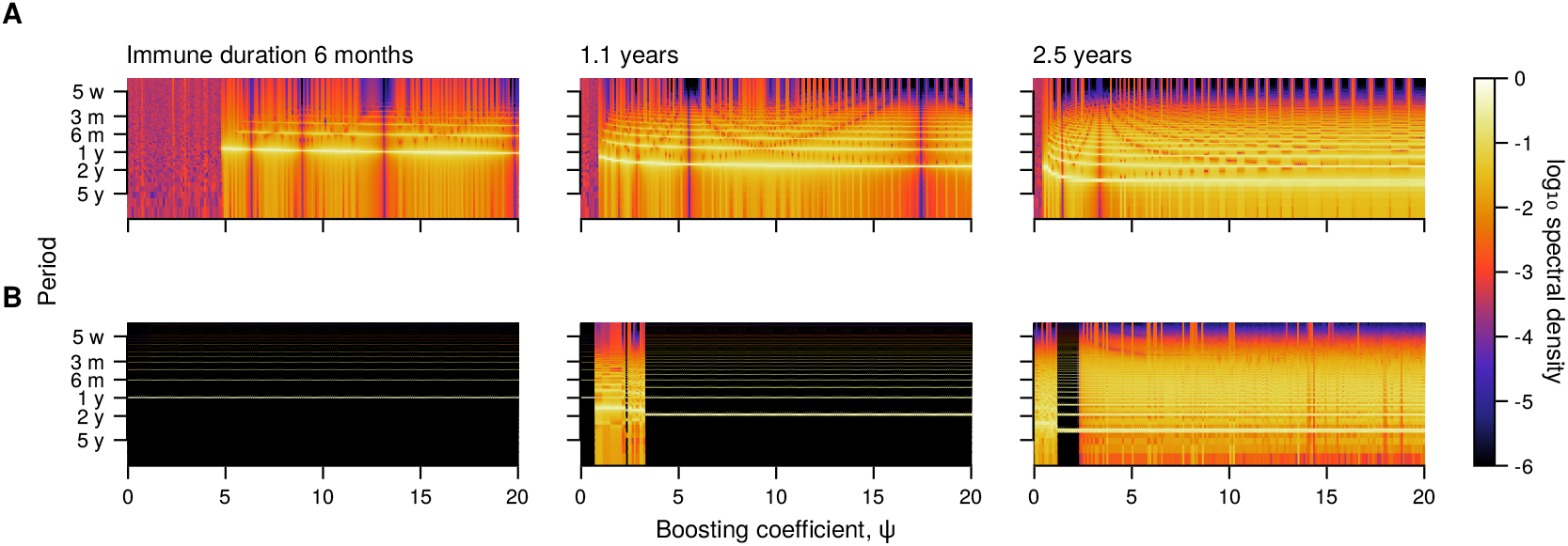
Periodicity of the model. (A) Discrete Fourier transform of simulations with constant parameters and (B) with seasonally forced parameters (amplitude 10% of mean transmission parameter). Bright areas show the period of outbreaks. In each plot, [inline], birth rate is 0.087 per year, and mean generation time is 7.5 days.

**Figure S5:**
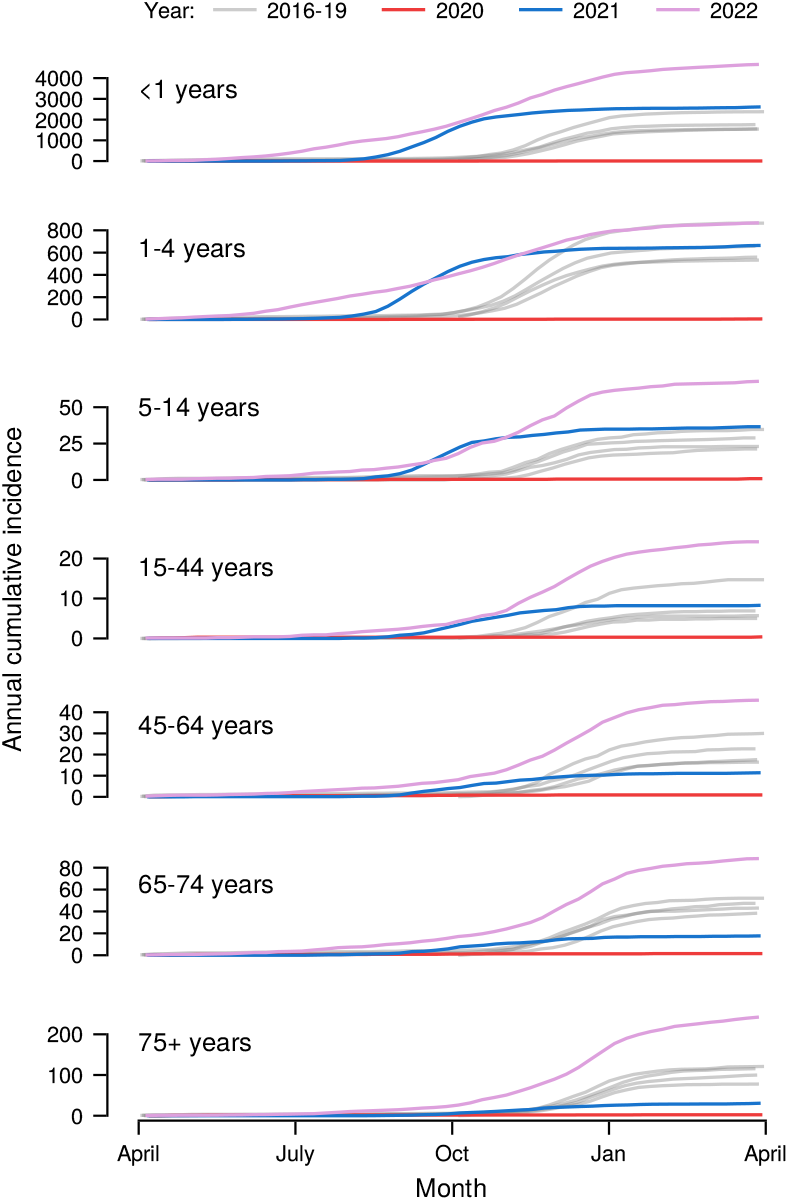
Age-stratified incidence of respiratory syncytial virus in Scotland. Each line represents cumulative number of recorded cases since the previous April.

**Figure S6:**
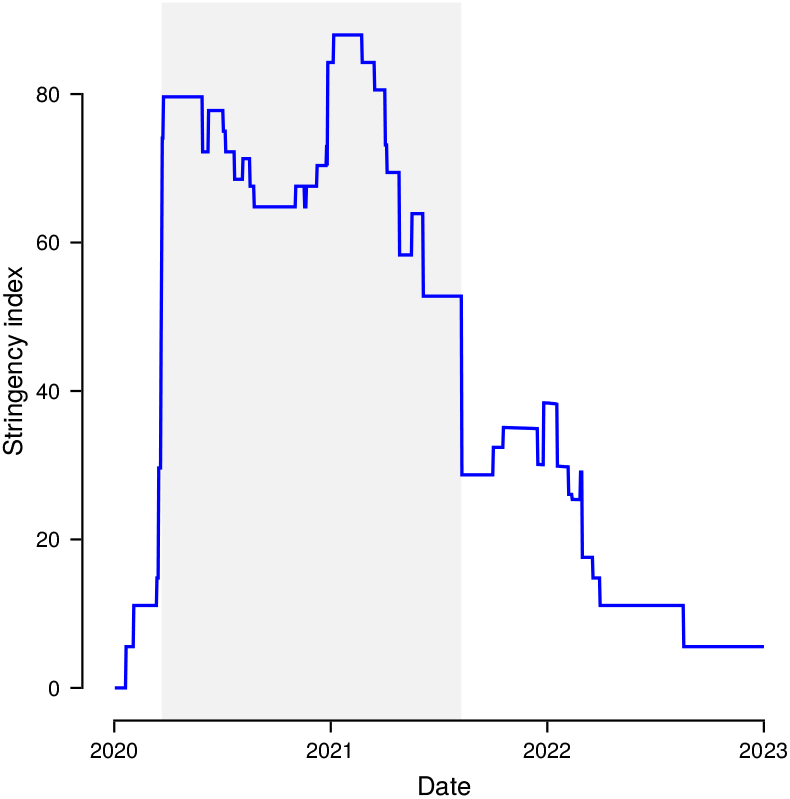
Stringency index in Scotland. The grey shaded area indicates the period when the stringency index was greater than 50. Data from Hale, et al., Nat *Hum Behav* **5**: 529-538 (2021).

## Supplementary text

### Parameter identifiability

We followed the method described by Castro and de Boer [28]. We re-wrote Equations 1-3 from the main text as functionally-independent summands. We scaled all parameters and compartment sizes by unknown scaling factors, *u*,

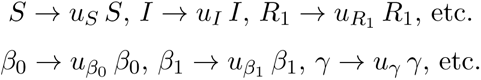

We assumed that prevalence was known without error, that is, *I(t)* was observed. This set *uI* = 1. We then equated each equation to its scaled version:

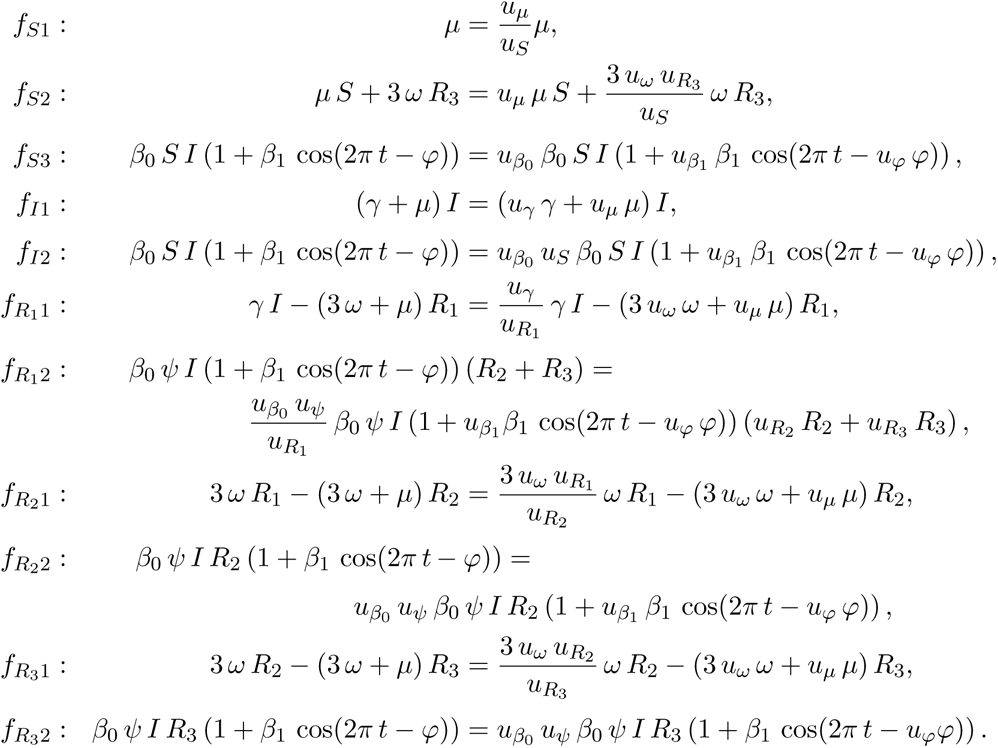

Rearranging these,

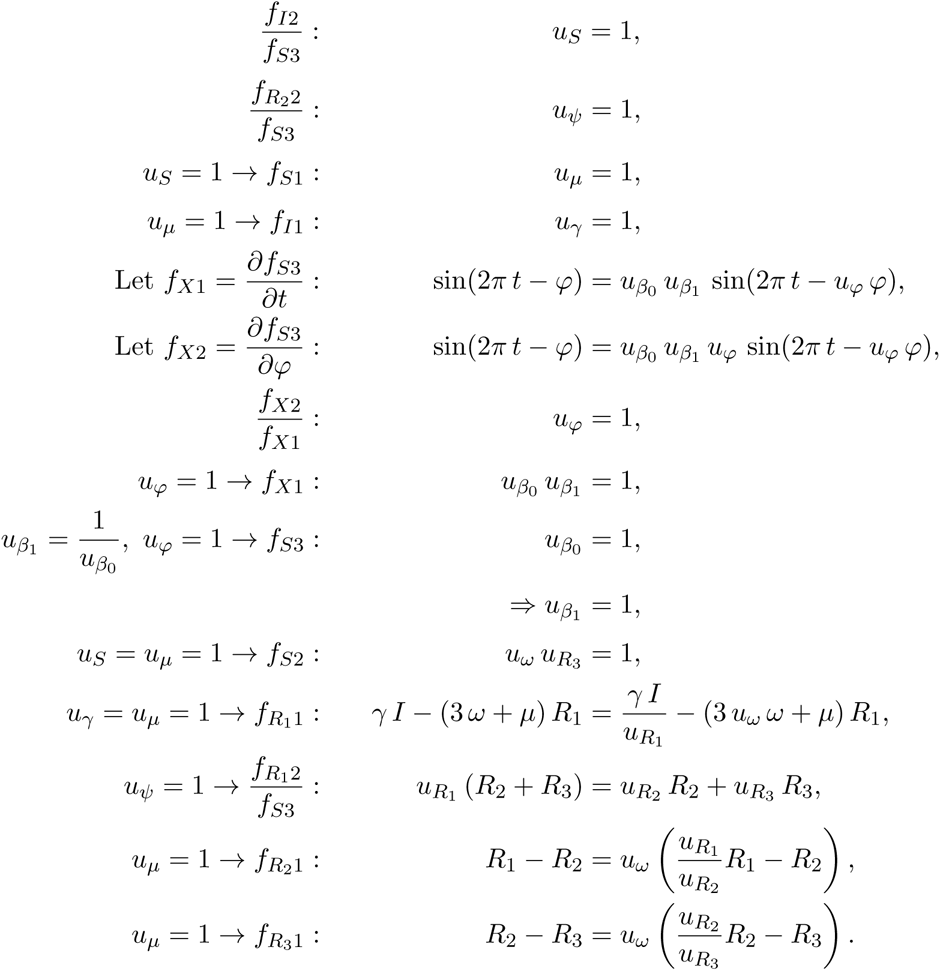

As these equations allow scaling parameters *u* ≠ 1, this model is not identifiable from prevalence data. To identify the parameters of this model, we would also need either to know the waning rate or be able to observe the size of one of the recovered subcompart­ments.

## Notes

### Competing Interest Statement

The authors have declared no competing interest.

### Author Declarations

Data used in this analysis were released by Public Health Scotland under the Open Government Licence for public sector information, https://www.nationalarchives.gov.uk/doc/open-government-licence/version/3/ available at https://www.opendat a.nhs.scot/dataset/48a9310e-4db3-44a4-bce8-6b4be9deb88a/resource/3 7beac86- f8fb- 4ab5- 9457- 2b8ddac9c089/download/respiratory_scot.csv

